# The monocyte-to-lymphocyte ratio: defining a normal range, sex-specific differences in the tuberculosis disease spectrum and diagnostic indices

**DOI:** 10.1101/2021.02.25.21251823

**Authors:** Thomas S. Buttle, Claire Y. Hummerstone, Thippeswamy Billahalli, Richard J. B. Ward, Korina E. Barnes, Natalie J. Marshall, Viktoria C. Spong, Graham H. Bothamley

## Abstract

**Background:** The monocyte-to-lymphocyte ratio has been advocated as a biomarker in tuberculosis. Our objective was to evaluate its clinical role in diagnosis, prognosis and treatment outcome.

**Methods:** Complete blood counts from an unselected population aged 16 to 65 years defined normal values of the ratio and associations with other indices. Blood counts, inflammatory markers and clinical parameters were measured in patients with and those screened for tuberculosis. We examined the ratio for its associations with these variables and for diagnosis, screening, prediction of poor prognosis and response to treatment. Results. In the unselected population, monocyte-to-lymphocyte ratios were higher in males than females and correlated with neutrophil counts (Spearman’s rho=0.48, P<0.00001, n=14,573). In 356 patients notified with tuberculosis, ratios were higher in males (high monocyte counts), especially in smear-positive pulmonary tuberculosis (S+PTB), lung cavitation and raised inflammatory markers. The sensitivities for confirmed tuberculosis were 42% (males) and 32% (females), with specificities of 70% and 71% respectively. Using sex-specific cut-offs in 629 adults screened for tuberculosis and with a positive tuberculin skin test or interferon-gamma release assay, diagnostic sensitivities for active tuberculosis were better in males (25%; all and contacts of a S+PTB index, respectively) than females (14-17%) with specificities of 89-96%. Positive likelihood ratios were better with upper limits alone but were still poor (6.64 when screening for tuberculosis, with an area under the curve of 0.688). Ratios did not predict death or response to treatment. Ratios were especially higher in males than female ratios in the 16-45 years age group.

**Conclusions:** Severe tuberculosis and male sex associated with high monocyte-to-lymphocyte ratios. The ratio performed poorly as a clinical aid. (269 words)

## Introduction

Tuberculosis (TB) was declared a global health emergency by the World Health Organization in April 1993 and remains a leading cause of death and disease [1]. The microbiological diagnosis of TB has advanced considerably through molecular testing [2]. The use of two proteins found in *Mycobacterium tuberculosis* but not in Bacille Calmette-Guérin (BCG) in interferon-gamma release assays (IGRAs) has improved the specificity of diagnosing latent tuberculosis infection (LTBI) [3].

Many non-specific biomarkers have been proposed to identify those with a positive IGRA who will develop active TB [4], those with sub-clinical disease (i.e. those without symptoms often with a normal chest x-ray without raised inflammatory markers) [5], those likely to die from TB [6], those whose genotype might suggest a protective response to *Mycobacterium tuberculosis* [7] and transcriptomic measures of risk, diagnosis and treatment response [8]. The ML ratio has experienced a revived interest as a biomarker. Originally identified in TB patients, a rabbit model suggested that both high and low values might be markers for TB progression [9]. In a mouse model, high ML ratios were associated with impaired protection of BCG against TB [10]. An elevated ML ratio was thought to correlate with different stages of TB disease [11], identify those with HIV infection most likely to develop active TB [12, 13], contacts likely to develop active TB [14] and neonates at risk of TB [15]. *In vitro* studies with a mycobacterial growth inhibition assay suggested that monocytes and lymphocytes from those with a higher ML ratio were less able to inhibit the growth of BCG [16]. We, therefore, examined the ML ratio in patients with TB and LTBI in order to test the hypothesis that such a simple measurement from a complete blood count could be of clinical value. Early results were reported in the form of abstracts [17, 18].

## Methods

### Study design

A retrospective audit of the value of ML ratios in patients with or screened for tuberculosis. Research was conducted according to the principles expressed in the Declaration of Helsinki.

### Setting

A university district general hospital in an inner-city London borough with a high incidence (62 to 22 per 100,000 during the period of study) of TB in a multi-ethnic population.

### Participants

Patients were eligible if they attended TB Clinics, had a complete blood count and attended between 2005 and 2018 (Figure 1). Children under 16 years of age were excluded. Active TB was defined by a) a positive culture or by b) a combination of consistent radiology, histology and other tests sufficient for the clinician to notify the patient as having TB and to start a course of treatment without a change in this opinion during treatment and within the following three years. Contacts were defined by their index case disease type, had spent > 8 h in close proximity to the index case, had attended the TB screening, had a positive IGRA or tuberculin skin test and were referred for consideration of preventive treatment. LTBI was defined in the screened populations as having a positive IGRA (> 0.35 IU/mL) or a tuberculin skin test (TST) ≥ 15 mm for those with a BCG scar and ≥10 mm for those without a BCG, and without evidence of active TB. All participants constituted a consecutive series (Figure 1).

**Figure 1.**
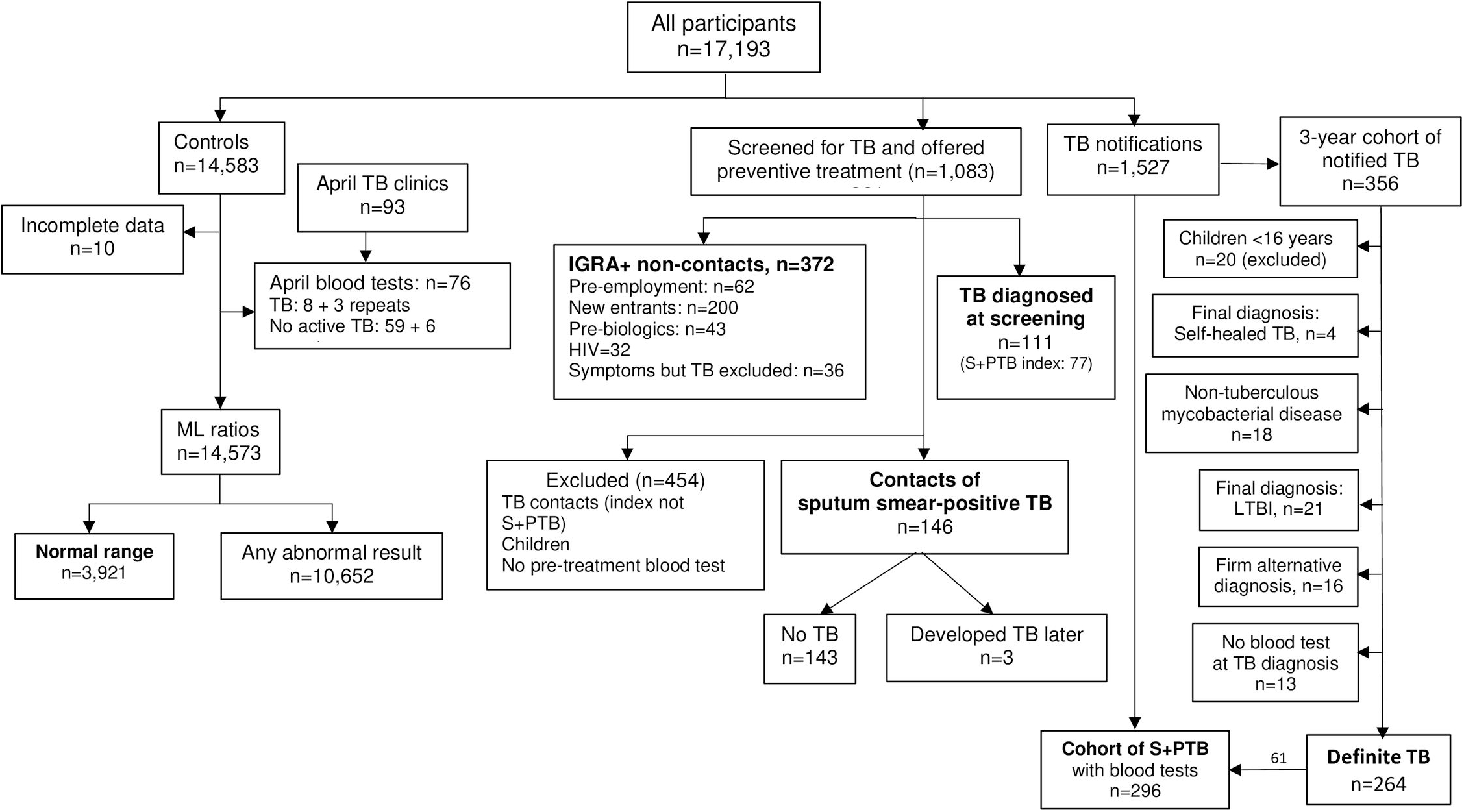
Study populations.

For demographic details, see Table 1 and Supplementary Table 2.

**Table 1.**
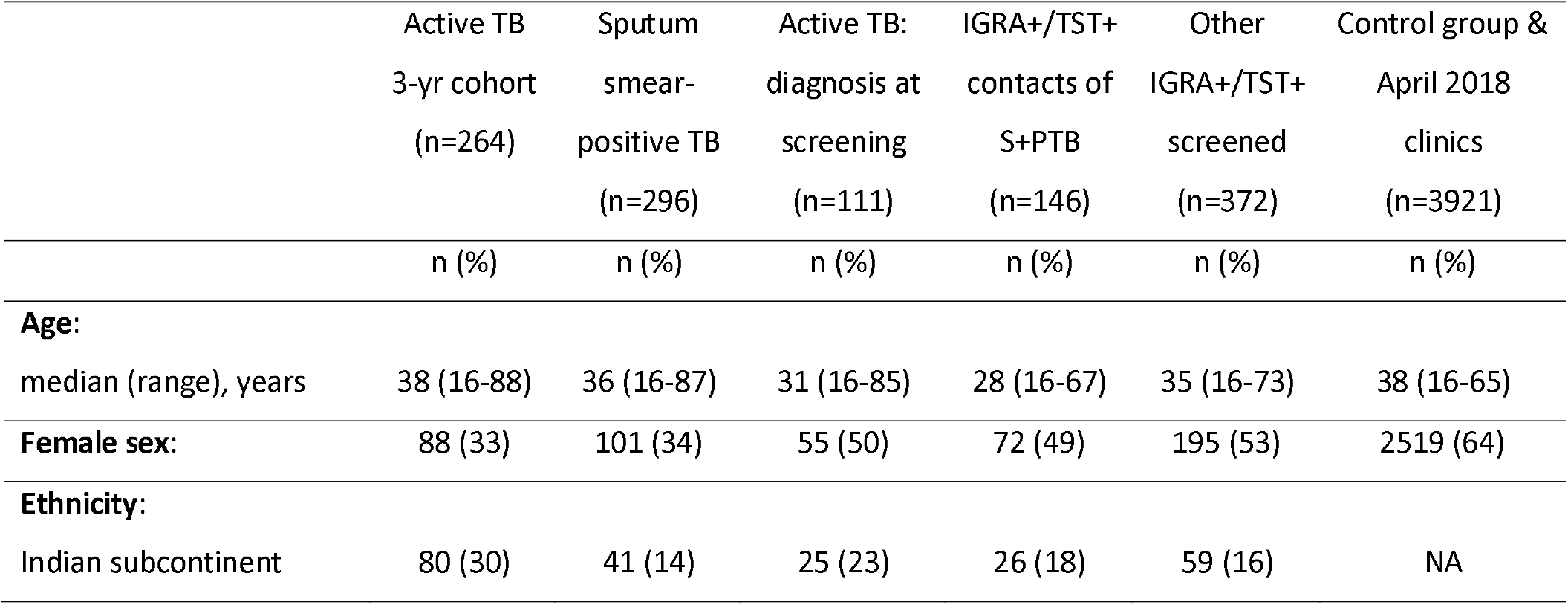
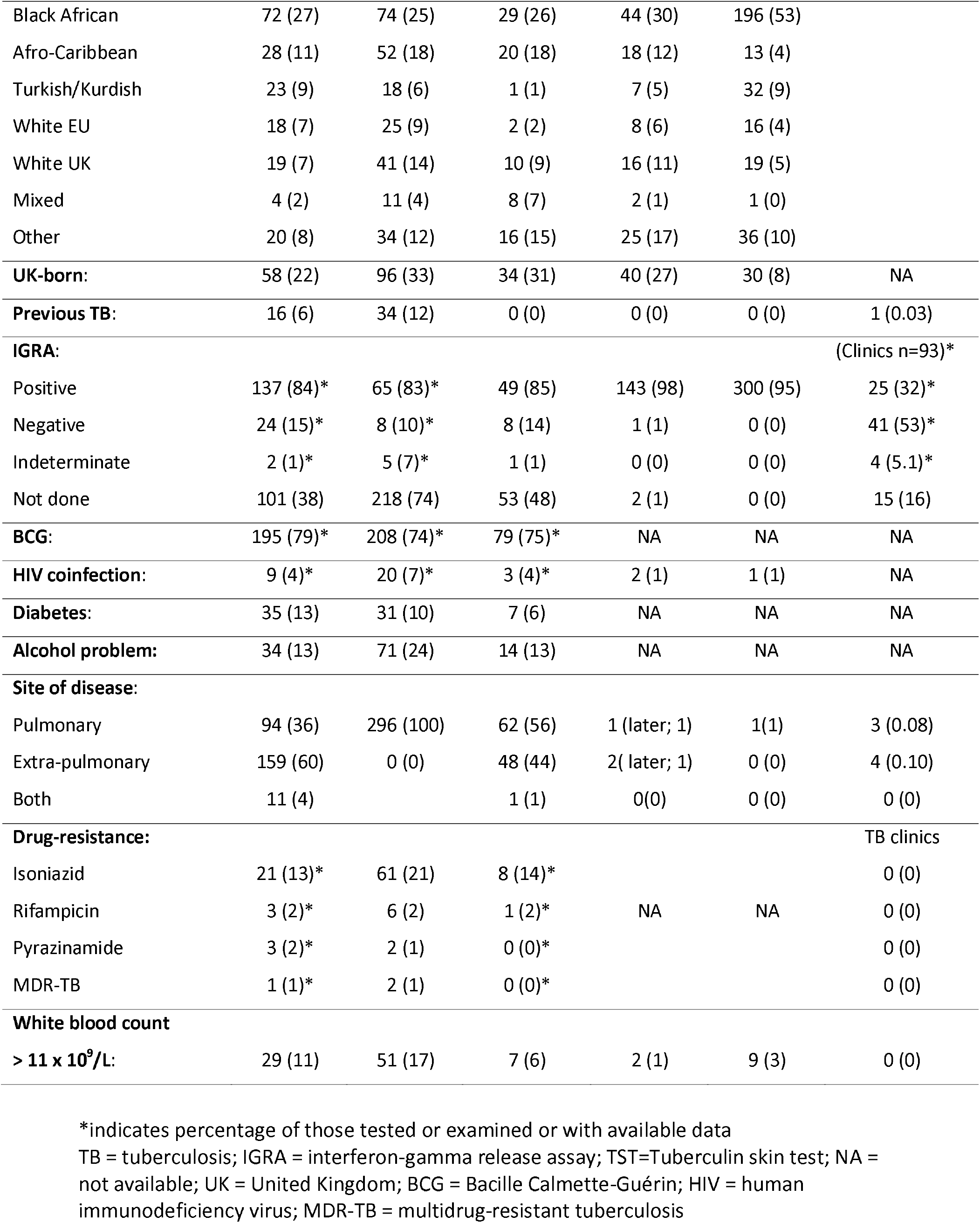
Demographic data

A control group was derived from anonymized complete blood counts analyzed during April 2018, excluding those over 65 years on the grounds of likely co-morbidities. Only data where the hemoglobin, total red cell count, packed cell volume, mean cell volume, mean cell hemoglobin, platelet count, total white cell count, neutrophils, lymphocytes, monocytes, eosinophils, and basophils were all within normal ranges were used to define a “normal range” for the ML ratio (Supplementary Table 1). Cut-off values were identified using the mean ± 2 SD from the normal data. An additional control group was derived from those notified as a case of tuberculosis where the diagnosis of active tuberculosis was subsequently excluded (see Figure 1).

### Variables

TB notification data (age, sex (self-identified), ethnicity (self-identified), country of birth, previous TB, BCG scar, HIV status, diabetes, problem alcohol use, site of disease (patient or index)) were recorded as required by Public Health England for TB as a notifiable disease. Chest radiograph zones out of six and cavitation, sputum smear, days to culture, drug susceptibility test (DST) results, sputum culture at 2 months and end of treatment, and outcome were noted. Deaths were classified as being a) due to TB, b) TB contributed or c) unrelated to TB. At screening, reason for attendance, tuberculin skin test (TST) and IGRA results were recorded. Blood samples were taken before the start of any treatment, at 2 months and the end of treatment.

### Statistical analysis

Data were analyzed with GraphPad Prism version 7 and Microsoft Excel. Cut-off ML ratios above and below the 95^th^ centile of the control group defined diagnostic indices. Receiver operating curve (ROC) analysis addressed the optimal cut off ML ratio and area under the curve (AUC) values (John Hopkins web-based calculator for ROC curves). Chi-squared analysis assessed the significance of ML ratios. Variables with a normal distribution were analyzed by parametric tests (Students’ t-test, Pearson’s correlation), whilst those with non-normal distributions were described by median and range and analyzed with non-parametric tests (Spearman’s rank correlation). A power calculation showed that to detect a 10% difference between the sensitivities of the ML ratio with an 80% power at the 5% level required 199 patients in each group if the higher sensitivity were 20%, or 293 if 30%.

## Results

### The anonymized dataset

Using anonymized complete blood counts taken during April 2018 from those aged 16-65 years (n=14,573), the ML ratio showed a distribution with a right-sided skew, which was normalized by log transformation (Figure 2). There was no significant difference in ML ratio in relation to sex or age. The ML ratio increased according to the neutrophil count (S1 Figure, Spearman’s ρ=0.48, P<0.00001); those with an ML ratio of 0.45 corresponded to those with a neutrophil count of 8-8.9 × 10^9^/L.

**Figure 2.**
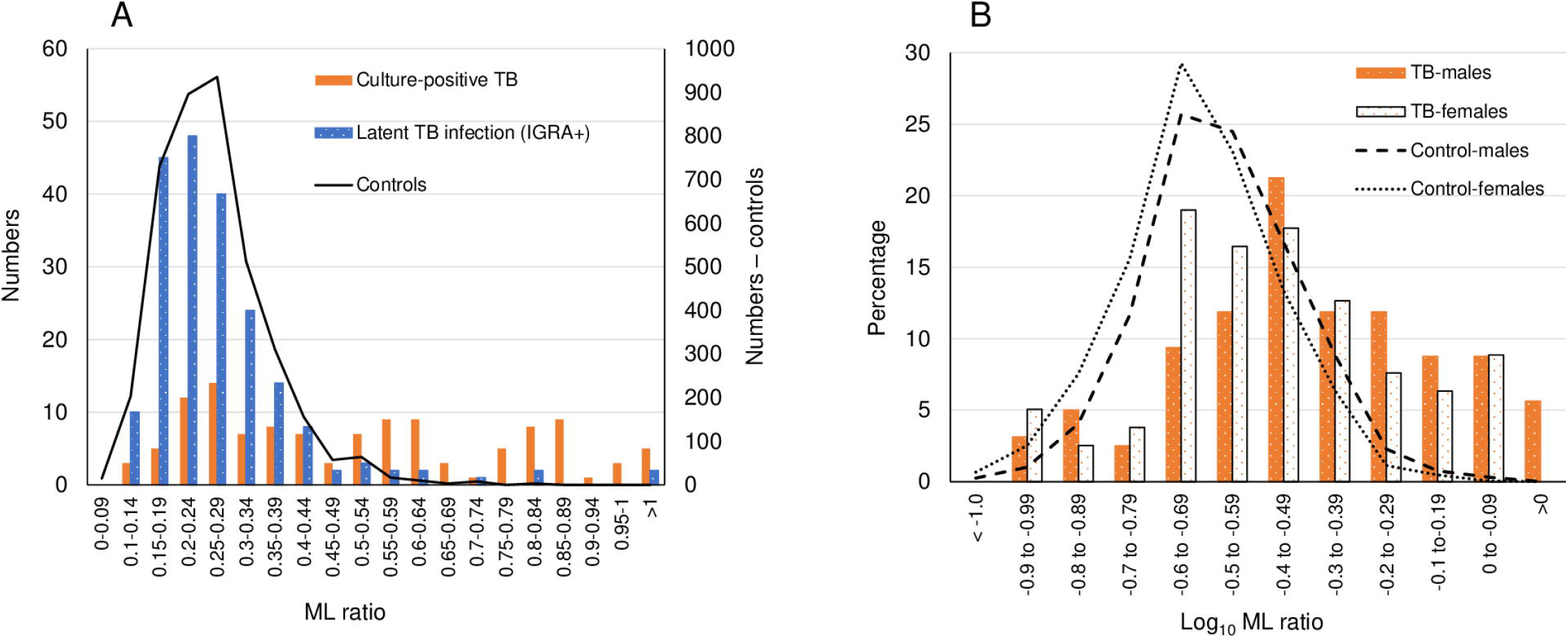
Monocyte-to-lymphocyte ratios and spectrum of tuberculosis (3-year cohort) **A**. Monocyte-to-lymphocyte ratios in patients with culture-positive tuberculosis (black columns) and screened individuals with a positive interferon-gamma release assay. This shows a right-sided skew of data. Latent tuberculosis infection (hatched columns) follows the control data (dash-dot line). **B**. Log-transformed monocyte-to-lymphocyte ratios in patients with tuberculosis related to sex. Normalized data indicates higher values for males in both controls (males - dashed line; females – dotted line) and tuberculosis (males - black stippled columns; females white stippled columns).

For the control group, complete blood counts with any abnormal value were deleted (n=10,652, 73% of the total dataset; Table 1 and S1 Table). More women than men had blood tests and so were more frequently represented in the control group (2519/3921 (64%)). ML ratios showed a right-sided skew (Figure 2A), which normalized with log transformation (Figure 2B). However, there was a significant difference in the log_10_ ML ratios between the sexes (mean (SD): female −0.619 (0.148); male, −0.582 (0.143), Student’s t-test, t=15.76). Defining cut-off values as log. mean ± 2SD, for females the upper ML ratio limit was 0.474 and for males was 0.505.

During April 2018, 8 patients with notified tuberculosis had 11 blood tests (Figure 1). In the various TB clinics, 93 individuals were seen, several on more than one occasion: none with LTBI or cured TB had an ML ratio above the upper limit for their sex, although one patient with sarcoidosis had an ML ratio below the lower limit. Those with a positive TST or IGRA did not have higher ML ratios than those who were negative.

### TB patients

356 patients were notified as having TB over the study period. Twenty were children and 13 had no pre-treatment blood test; 50 were subsequently denotified (Figure 1). The firm alternative diagnoses included cancer, pneumonia, abscesses, Crohn’s disease, glomerulonephritis, lupus and autoimmune encephalitis, non-tuberculous mycobacteria, self-healed tuberculosis and LTBI. Demographic details of the remaining 264 patients are given in Table 1.

Six patients had an ML ratio below the lower cut-off limits, two with culture-positive TB; two had diabetes, one HIV co-infection (culture-positive), one an alcohol problem, one with malnutrition (culture-positive) and one with no co-morbidities. More males with TB had an ML ratio > 0.505 than those below this limit, but the difference was not significant. Fewer patients from the Indian subcontinent had ML ratios above the upper limit of normal, but the difference was not significant. There were too few patients with previous TB, HIV infection, diabetes or an alcohol problem to determine any differences, but the percentages were similar in those above and below the sex-specific ML ratios.

More patients with culture-positive TB from any site had a high ML ratio (48/69 (70%) vs. 48/108 (24%); χ^2^ = 10.7, P = 0.001) compared to those without a positive culture. More patients with pulmonary disease had ML ratios above the upper limit compared to those with extrapulmonary disease (46/94 compared to 44/159; χ^2^ = 10.8, P = 0.001, see Figure 3). Three patients with miliary TB had ML ratios of 0.10 (lymphocytes 5.1 × 10^9^/L), 0.36 and 1.00 (bone marrow depression with lymphocytes and monocytes both with an absolute value of 0.2 × 10^9^/L). Three patients with problem alcohol use and disseminated TB had high ML ratios due to lymphopenia (all three had a lymphocyte count of 0.6 × 10^9^/L).

**Figure 3.**
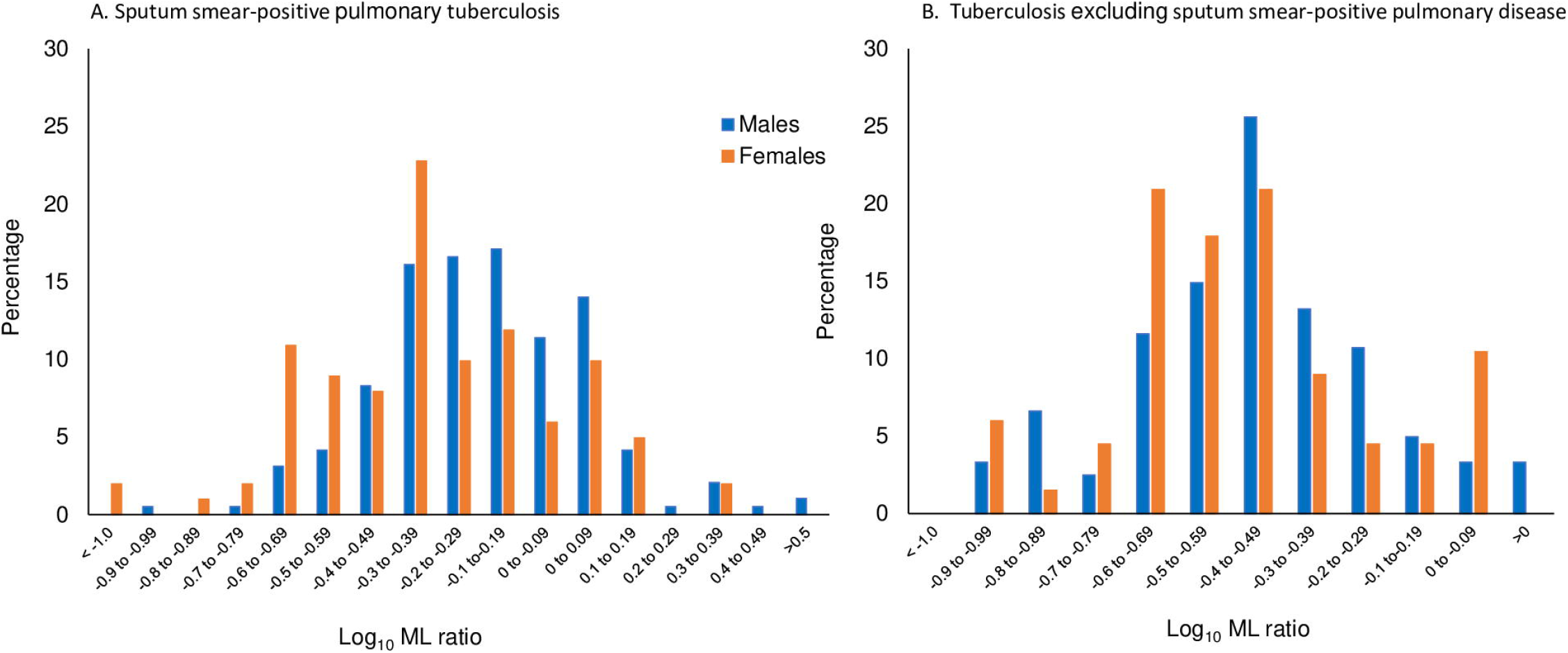
Monocyte-to-lymphocyte ratios and sex. **A**. Patients with tuberculosis excluding sputum smear-positive pulmonary disease. **B**. Patients with sputum smear-positive pulmonary tuberculosis. Males (blue) show higher values than females (orange).

White cell counts >11 × 10^9^/L were more common in those with ML ratios above their sex-specific limits (19/96 vs. 10/168; χ^2^ = 12.0, P = 0.0006). Albumin levels were more commonly <40 g/L in those with a high ML ratio (80/90 compared to 103/167; χ^2^ = 18.9, P = 0.00001) and globulin >32 g/L were higher (83/90 compared to 133/163; χ^2^ = 4.9, P = 0.027). C-reactive protein (CRP) levels were high (>10 mg/L) in those with ML ratios above the sex-specific limits (74/87 vs. 61/140: χ^2^ = 38.3, P < 0.00001) and higher in males with values >10 mg/L than females (Mann-Whitney, z=2.007, P = 0.045).

### Diagnostic indices (Table 2)

Screened persons who were IGRA+ were identified from records of those offered preventive treatment (Figure 1). Contacts of patients with S+PTB (n=146) were considered separately (Table 1) as being most likely to develop active disease as a result of recent infection. For more distant infection, only screened individuals with a positive IGRA were included (n=372).

**Table 2.** Sensitivity and specificity of sex-specific ML ratio cut-off values: mean ± 2SD healthy controls

| Population | Sex | No. | True positives | False positives | False negatives | True negatives | Sensitivity (%) | Specificity (%) |
| --- | --- | --- | --- | --- | --- | --- | --- | --- |
| **General CBCs <sup>a</sup>** |  |  |  |  |  |  |  |  |
|  | F | 9977 | 0 (0)<sup>b</sup> | 1552 (1297) | 2 (2) | 8423 (8678) | 0 (0) | 85 (87) |
|  | M | 4594 | 2 | 794 (686) | 7 | 3791 (3899) | 22 (22) | 83 (85) |
| **TB notifications <sup>c</sup>** |  |  |  |  |  |  |  |  |
|  | F | 112 | 28 (27) | 7 (5) | 60 (61) | 17 (19) | 32 (31) | 71 (79) |
|  | M | 203 | 74 (69) | 8 (7) | 102 (107) | 19 (20) | 42 (39) | 70 (74) |
| **All screened excluding contacts** |  |  |  |  |  |  |  |  |
|  | F | 251 | 8 (7) | 11 (5) | 48 (49) | 184 (190) | 14 (13) | 94 (98) |
|  | M | 233 | 14 (11) | 14 (4) | 42 (45) | 163 (173) | 25 (20) | 92 (98) |
| **Contacts of S+PTB index vs. S+PTB diagnosed after screening** |  |  |  |  |  |  |  |  |
|  | F | 106 | 6 (5) | 3 (2) | 30 (31) | 67 (68) | 17 (14) | 96 (97) |
|  | M | 117 | 11 (9) | 8 (3) | 33 (35) | 65 (70) | 25 (21) | 89 (96) |

CBC=complete blood count; F=female; M=male; TB = tuberculosis; S+PTB=sputum smear-positive pulmonary tuberculosis. <sup>a</sup> Sex for two samples not available. <sup>b</sup> Figures in brackets are for upper limits only. <sup>c</sup> 50 denotified cases included.

The ML ratio for those with LTBI, whether with recent or distant exposure, showed an almost normal distribution with data skewed towards higher ML values (Figure 2A). Log-transformation of the data showed a normal distribution, except for patients with TB (Figure 2B). Males with TB had more ML ratios above the upper limit than females (Table 2; χ^2^ = 10.1, P = 0.001). The positive likelihood values for a ML ratio were 1.38 in the April 2018 population, 2.92 in the screened population and 2.76 in contacts of S+PTB; if only high ML cut-off values were used, the positive likelihood ratios improved to 1.63, 6.64 and 8.01 respectively (Table 2) but were still <10. Assessing receiver-operator curves, the areas under the curve for high ML ratios, omitting the April 2018 population review, were 0.688 for the screened population (0.669 if denotified patients who were screened were included) and 0.698 for contacts of S+PTB, respectively.

### Prognostic value of the ML ratio in those with LTBI

In contacts of index cases with S+PTB, there was one patient with an ML ratio of 1.0 who on review was considered to have active TB, based on atelectasis in left lower zone, TST conversion from 6 to 15 mm induration and a CRP of 28 mg/L and whose chest radiograph cleared following treatment for TB (Table 2). Two other patient who developed culture-positive TB had ML ratio of 0.26 and 0.22 at screening.

There were 11 contacts of S+PTB who showed a completely negative response to tuberculin at their first visit with a positive TST at 6-8 weeks follow-up. Eight of nine with an IGRA were positive (one rising from 0 to 0.5 IU/mL). All these with evidence of recent infection with TB had an ML ratio within the normal ranges and none developed TB (all received preventive treatment).

In the IGRA+/TST+ screened population without known contact with TB, 8 had a high ML ratio (see Table 2). One had initially been notified as a case of TB being a contact of an uncle, who had active but not pulmonary TB; this person had several risk factors for TB, minor gastro-intestinal symptoms and was given a trial of standard treatment which was stopped at 2 months (2 months of rifampicin and pyrazinamide being a proven preventive treatment for LTBI). The other 7 did not develop TB, having received preventive treatment.

### Deaths and ML ratio

There were 29 deaths (1.9%) in notified TB cases from 2005-2017. Three were due to TB, three where TB contributed to death and the remainder were unrelated to TB. ML ratios above the sex-specific limits were found in 18 (62%) but were not significantly higher or more numerous in those where TB played a role (5 males and 1 female) compared to those where there was another cause of death. ML ratios measured within a day or two of death did not improve on the predictive value above samples taken at the time of TB diagnosis.

### ML ratios in smear-positive pulmonary tuberculosis (S+PTB)

Preliminary examination of patients with S+PTB from the 3-year cohort had shown possible differences between those with a high ML ratio and those within the normal range [18]. In order to resolve the possibility of type II errors, the sample with S+PTB included all such patients over a 12-year period (Figure 1; see also S2 Table).

The female-to-male ratio in S+PTB was 1 to 2.0. There were no differences in ML ratios related to demographic features, but higher ratios were still more frequent in males (130/194 cf. 56/101; χ^2^ = 3.81, P = 0.05). Those with a high ML ratio were more likely to have a sputum smear of 3+ (χ^2^ = 9.0, P = 0.0027), but the difference between males and females was not significant. There was no difference in the radiographic extent of disease, but cavitation was more frequent in females with high compared to normal ML ratios (χ^2^ = 10.0, P = 0.0016) and in males than females with normal ML ratios (χ^2^ = 9.6, P = 0.00195).

The ML ratio did not predict culture status at 2 months, the duration of treatment nor the presence of isoniazid resistance. Albumin levels were lower (Students’ t-test: t=5.3, P<0.0001, df 288), globulin levels were higher (t=3.52, P=0.0005) and CRP values were higher (Mann-Whitney U-test P<0.00001) in those with a high ML ratio at the time of diagnosis; there was no difference between males and females for these biomarkers.

Two-week follow-up ML ratios were available for 178 with S+PTB. Five showed no change, 57 were higher and 115 were lower; ML ratios were more likely in males than females to fall to the normal range (χ^2^ = 6.5, P = 0.011). Looking only at results obtained at the start of treatment, at 2 months and at the end of treatment, there was a significant fall in the ML ratio by 2 months (paired t-test: t=7.5, P < 0.0001, df 136) and 6 months (t=5.3, P<0.0001, df 117). Looking at individual patients, a persistently high ML could not be used to predict who would remain culture-positive at 2 months or have a poor outcome. Four patients had extended inpatient stays and multiple blood tests taken due to their severity of disease. Three showed a reduction in ML ratio from high to normal values by days 2, 6 and 21 respectively, whilst another showed an ML ratio which remained high for 247 days.

In order to determine whether the sex difference in ML ratios might be due to sex per se, sex hormones or differences in inflammatory responses with age ML ratios were divided by age groups 16-45 years, 46-65 years and >65 years. In order to ensure that the level of disease burden was comparable, only those with S+PTB were examined. The 16-45 years age group showed the greatest difference in ML ratio between the sexes (Student’s t-test: t = 2.96, P = 0.0035), although the 46-65 years age group was also significantly different (t = 2.03, P=0.047; Figure 4).

**Figure 4.**
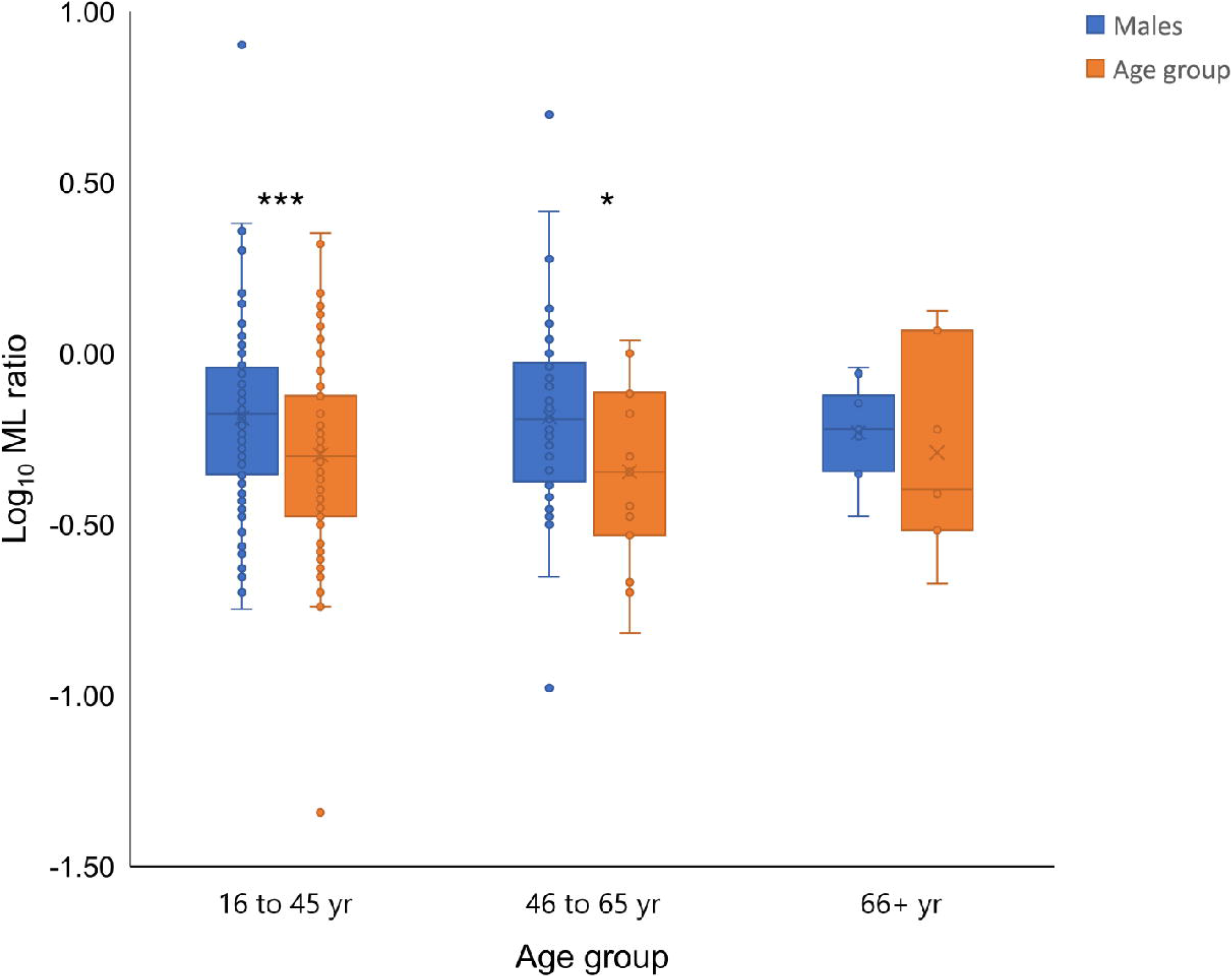
Age and sex differences in monocyte-to-lymphocyte ratios.

ML ratios were compared using log-transformed values by Student’s t-test. * P < 0.05, ** P < 0.01 and *** P < 0.005. Mean ML values were 0.65 for males in the 16-45 and 46-65 age groups and 0.50 and 0.45in females, respectively.

## Discussion

In an unselected population, ML ratios correlated with neutrophil counts. High ML ratios were more often due to monocytes, especially in males. The sensitivity of ML ratios was poor for the diagnosis of TB or as a screening tool to detect active TB in contacts and IGRA+ individuals. High ML ratios did not predict the development of TB in screened populations, nor treatment effectiveness, although ML ratios fell with treatment. However, high ML ratios were associated with pulmonary compared to extrapulmonary TB and markers of systemic inflammation (neutrophil count, low albumin, high globulin and high CRP values), especially in males. In S+PTB, high ML values were associated with sputum smears of 3+ and lung cavitation, but not with radiographic extent of disease. The sex difference was most marked in the 16-45 years age group.

### Limitations

This audit was performed at a single site. However, this ensured that data collection was uniform and follow-up data consistently available. Few patients had HIV co-infection and those with HIV were taking antiretroviral therapy and had normal lymphocyte counts. Children were excluded, but this avoided a bias in terms of empirical treatment for TB and a population in which blood tests are fewer and for highly selected reasons. Those over 65 years of age, excluded in the control group, are likely to have disease co-morbidities, especially malignancy, ischemic heart disease and metabolic syndrome [19, 20], which are associated with higher ML ratios. Older persons also experience “inflammaging”, having a higher inflammatory response than younger adults [21-23].

### Sex-specific differences

Males had higher ML ratios than females. Males have more S+PTB, whereas the sex ratio in extrapulmonary TB is equal [24, 25]. Lung cavities are more common in males than females [26]. Many of the reported data on ML ratios either had a predominance of males in the TB group [11, 27] or did not give sex-specific data [16]. The immune response in males is less effective than in females despite greater TLR2 and TLR4 expression, more Th17 cells and CD4+ Treg cells, CD8+ T cells and NK cells [28]. Monocytosis is a feature of infection [29] and high values are found especially in males [30]. Thus, the higher ML ratios in males and their association with more severe forms of TB and with lung cavities might be expected as a sex-specific immune response. Such individuals are more readily diagnosed and often bias studies in TB towards a male-predominant population.

### Clinical value

The early use of the ML ratio in TB noted that “the … ratio cannot … be used as a criterion for diagnosis”, the “monocyte-lymphocyte ratio … is not a sensitive index of activity in the tuberculous patient” (activity being measured by the extent of disease, tachycardia, fever, clinical judgment of prognosis and follow-up over two years) and that “a considerable number of patients with tuberculosis will have a normal monocyte-lymphocyte ratio” [31]. These results from the pre-antibiotic era of TB treatment have been confirmed in a modern setting by this study. The association of lymphopenia and a lower ML ratio with TB in a rabbit model [9, 32] was rarely observed in our study, even in those with miliary disease as the human comparator of the experimental method.

A low or high ML ratio has been associated with a hazard ratio of 2.47, 1.5 and 1.22 for the development of TB during follow-up of adults with HIV infection before anti-retroviral therapy (12), in HIV-infected post-partum women (13) and in infants (15) in South Africa, respectively. However, the sensitivities of cut-off values were low (< 10%), such that the ML ratio had little predictive value for an individual person. A higher hazard ratio (4.5) was observed in contacts who went on to develop TB, but a combination of a tuberculin skin test ≥ 14 mm (sensitivity 7.5%) with monocytes >7.5% of all white blood cells (sensitivity and specificity about 75%) was reported to have a hazard ratio of 8.78, noting that many of the diagnoses of TB were a decision to treat in young children (14). Using the same criteria from previous studies in our group of contacts, the sensitivity and specificity were poorer than in previous studies and the positive predictive value was low (Supplementary Tables 3 and 4).

### Generalizability

The use of a real-life group of TB patients and contacts from many ethnic backgrounds encourages extrapolation to the global HIV-uninfected TB population. Analysis of groups by smear and culture status, ensured that both real-life results and data uncontaminated by variations in the diagnosis of “clinical TB” could be obtained.

### Implications

This study has shown the importance of sex as a variable in the evaluation of biomarkers. Such differences touch upon lipid and metabolic differences in cardiovascular disease [33], cancer biomarkers [34] and studies distinguishing general differences between the proteome [35] and transcriptome [36, 37] between the sexes. Both estrogens and androgens affect immune responses [28, 38]. Our data suggested that the difference in ML ratios might be due to sex hormones rather than an X-linked cause or sex-related differences in inflammaging. Thus, diseases with a female or male predominance, such as autoimmune diseases or cancer respectively, may need to address the relative efficacy of diagnostic markers and prognostic indicators between the sexes.

Biomarkers and the sexes are especially problematic in tuberculosis. The male predominance is found almost entirely in S+PTB [24, 25] and therefore patient selection will be affected by the number with this form of disease. The relative ease of diagnosis of this form of TB again means that follow-up studies of, say, those with LTBI who go on to develop TB will mean that prognostic biomarkers will be biased towards males.

For example, transcriptomic data investigating the pathogenesis of active TB disease show sex-specific responses [39, 40]. An IL6/IL6R/CEBP gene module has been associated with monocyte expansion, high ML ratios, high CRP levels and a positive sputum smear [41]. The link to this module was closer than to a group of transcripts with a shared IL6-Type I interferon gene module associated with a high ML ratio per se [16, 41]. Estrogens, via the alpha receptor, downgrade IL6 expression in human monocytes [42]. The 3-gene transcriptomic signature of Sweeney, which performed better than 15 other signatures for discriminating latent from active TB [43], has genes which are affected by sex. Dual-specificity phosphatase 3 (DUSP3) deletion protects female but not male mice from endotoxin shock due to a dominance of M2 (non-classical and anti-inflammatory) macrophages [44]. Guanylate-binding protein 5 (GBP5) is a marker of IFNγ-induced classical monocytes [45], which are more frequent in males, and has been found in those who develop innate immunity after an IFNγ response [46]. Kruppel-like factor 2 (KLF-2), in association with the male-dominant cytokines interleukin-6 and monocyte chemoattractant protein-1, promotes inflammation in response to tumor necrosis factor (induced by mycobacterial lipoarabinomannan) [47]. If biomarker studies select a majority of males and/or those with S+PTB, their value might be less in females and those with more limited disease.

### Conclusion

The ML ratio showed a bias towards males with S+PTB, a fact which might account for its association with S+PTB and its use as a biomarker in tuberculosis.

## Supporting information

Data dictionary for column headings

Data from study participants

Anonymised control data

## Data Availability

Anonymised data is available in the data supplements.

## Acknowledgements

We thank Bala Sirigireddy for providing the hematology data used to define the normal ranges of the monocyte-to-lymphocyte ratio.

## Notes

### Competing Interest Statement

The authors have declared no competing interest.

### Clinical Trial

The study was an audit

### Funding Statement

This work was supported by the Homerton University Hospital Research and Development Office/North Thames Clinical Research Network (NIHR 4177; TSB 0.5 wte salary), but all other authors received no specific funding for this work.

### Author Declarations

Blood Tests in Tuberculosis III was approved by the East London and City Health Authority Research Ethics Committee (P/03/285), registered under the number NIHR 4177. The UK PREDICT TB procedures and protocol were approved by the Brent NHS Research Ethics Committee (10/H7017/14). Both gave access to the data for full blood counts, which were used in this study. Waivers for the full blood count data were given for those who were screened for participation in these studies.

