## Supplementary figures and images for "The monocyte-to-lymphocyte ratio: defining a normal range, sex-specific differences in the tuberculosis disease spectrum and diagnostic indices"

### Supplementary Figure

#### Slide 1
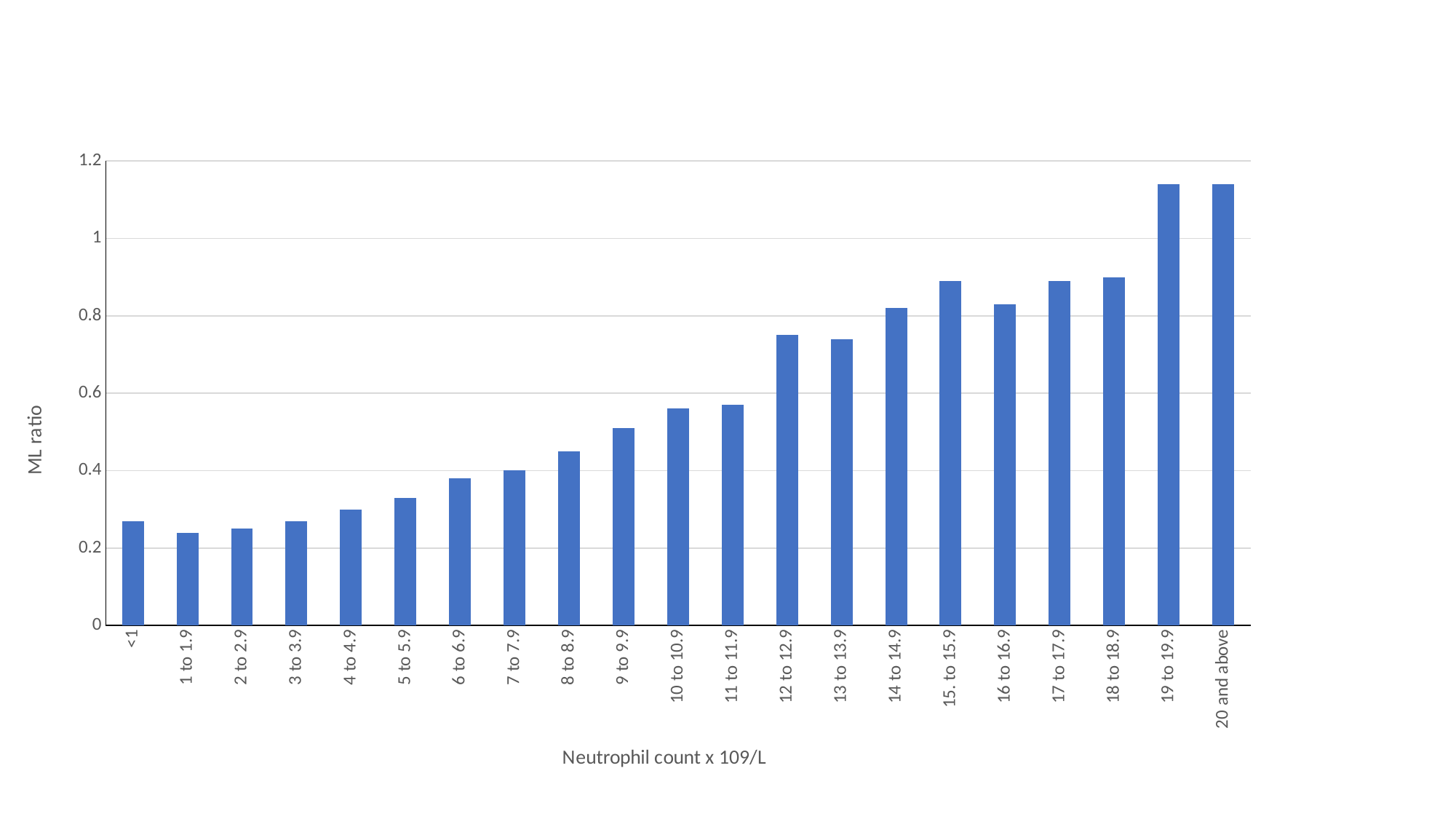

##### Chart
| Category | mean ML ratio |
|---|---|
| <1 | 0.27 |
| 1 to 1.9 | 0.24 |
| 2 to 2.9 | 0.25 |
| 3 to 3.9 | 0.27 |
| 4 to 4.9 | 0.3 |
| 5 to 5.9 | 0.33 |
| 6 to 6.9 | 0.38 |
| 7 to 7.9 | 0.4 |
| 8 to 8.9 | 0.45 |
| 9 to 9.9 | 0.51 |
| 10 to 10.9 | 0.56 |
| 11 to 11.9 | 0.57 |
| 12 to 12.9 | 0.75 |
| 13 to 13.9 | 0.74 |
| 14 to 14.9 | 0.82 |
| 15. to 15.9 | 0.89 |
| 16 to 16.9 | 0.83 |
| 17 to 17.9 | 0.89 |
| 18 to 18.9 | 0.9 |
| 19 to 19.9 | 1.14 |
| 20 and above | 1.14 |
